# Participant Demographics and Testing Trends: A Community Pandemic Response Program

**DOI:** 10.1101/2023.01.02.23284119

**Authors:** Yury E. García, Leslie Solis, Maria L. Daza–Torres, Cricelio Montesinos-López, Brad H. Pollock, Miriam Nuño

## Abstract

**Objective:** This study aimed to identify demographic characteristics of test participants and changes in testing participation over time in a community pandemic-response program launched in a college town in California, USA.

**Methods:** We described overall testing participation, identified demographic characteristics of frequent testers, and evaluated changes in testing participation over four different periods of the COVID-19 pandemic.

**Results:** A total of 770,165 tests were performed between November 18, 2020, and June 30, 2022, among 89,924 residents of Yolo County (41.1% of population), with significant participation from racially/ethnically diverse participants and across age groups. Most positive cases (49.9%) were captured during Omicron, which also corresponds to the period with the highest daily participation (895 per 100K population). The proportion of participants which we considered “frequent testers” (28.9% vs. 39.7%, *p <* 0.0001) and individuals that tested once (39.5% vs. 47.9%, *p <* 0.0001) increased significantly from Delta to Omicron. Women (58.8%), participants of age 19-34 years (38.8%), and White (53.2%) tested more frequently throughout the program. The proportion of tests conducted among Latinos remained steady around 18% over time, with the exception of the post-Omicron period (13%).

**Conclusion:** The unique features of a pandemic response program that supported communitywide access to free asymptomatic testing provides a unique opportunity to evaluate adherence to testing recommendations, and testing trends over time. Identification of individual and group-level factors associated with testing behaviors is essential to improve access and protect communities at-large.

## 1 Introduction

The COVID-19 pandemic presented major challenges and demanded effective public health responses that involved sustained behavior change, stay-at-home restrictions, utilization of face coverings, testing, and vaccination. Public health guidelines are a critical first line of a pandemic response but rely on compliance with evolving recommendations and restrictions. Trust and compliance are driven by complex factors, including individuals’ beliefs [1], risk perception [2, 3], trust in government [4], and demographic factors [5, 6, 7] that may vary throughout a pandemic, making containment particularly challenging.

Widespread testing and timely diagnosis are critical for pandemic control and preparedness. Mass testing was one of the essential control measures for curtailing the burden of the COVID-19 pandemic, particularly during its early phases [8]. Early detection of cases can limit viral transmission by isolation, quarantine, and contact tracing [9, 10, 11]. Over the course of the pandemic, new preventive and surveillance mechanism surged, including vaccine programs, wastewater surveillance [12], and at-home COVID-19 tests [13]. The virus itself and protective behaviors also changed throughout the waves of the pandemic [1]. Government and public health officials implemented health contingencies that were adaptable and flexible to reduce socioeconomic and long-term health burden fatigue [14, 15].

Understanding how people’s adherence to preventive measures changes over time has the potential to guide policy-makers as they amend strategies to revitalize public health strategies in future outbreaks. Studies of previous natural disasters suggest that people’s perceptions of risk and their responses to such risk may vary between individuals. Gender, age, socioeconomic status, personal experience of a natural hazard, and trust in authorities are factors that affects people’s responses to catastrophic events [16, 17, 18]. Women are more prone to panic during a crisis; they also report greater fear of disasters and perceived risks than males [19]. The COVID-19 pandemic is no exception. A growing body of research reported factors associated with adherence and non-adherence to COVID-19 preventive measures [20, 21, 1, 2, 22], namely, age [5], gender [6, 7], higher education level [23], marital status, and having children [24].

Healthy Davis Together (*HDT*) [25] is a program that was implemented in the City of Davis (California), as an effort to mitigate the spread of COVID-19 and facilitate the return to normalcy. Launched on September 2020, *HDT* focused on curtailing the burden of the pandemic among residents and workers in Davis. On July of 2021, the program was expanded to support residents of Yolo County in a re-branded effort, Healthy Yolo Together (*HYT*). The program consisted of numerous simultaneous interventions. First, it provided access to free saliva-based asymptomatic testing with high throughput methods to process large volumes of tests. It promoted targeted communication campaigns to improve participation in testing, provide access to isolation protocols for individuals sick and/or exposed. Education efforts were implemented to increase awareness and reduce misinformation; large-scale distribution of personal protective equipment; partnerships with the community to foster trust; and investments in the local economy to help businesses and workers.

This study aims to provide a detailed description of tests conducted by HDT with a focus on participant characteristics and notable changes in test participation over time.

## 2 Methods

### 2.1 Data description

*HDT* efforts to provide free asymptomatic testing expanded beyond the City of Davis to residents of Yolo County and California state-wide. For the analysis presented in this study, we will summarize tests conducted in residents of Yolo County. This assumption was made to ensure robust estimates of population-level participation in testing using the 2020 Census Bureau Survey [26]; we assume a total of 218,774 people in Yolo County and 68,640 in the City of Davis. A total of 770,165 tests were conducted in Yolo County among 89,924 people, of whom 53,869 reside in the City of Davis. Data is presented according to relevant periods throughout the pandemic: *Pre-Delta* (11/18/20–6/13/21), *Delta* (6/14/21–12/20/21), *Omicron* (12/21/21–3/15/22), and *Post-Omicron* (3/16/22–6/30/22). These periods were defined according to the dominance of a variant as reported by the California Department of Public Health [27, 28] (Figure 1). Delta B.1.617.2 and Omicron B.1.1.529 variants – hereafter referred to as *Delta* and *Omicron*, respectively.

**Figure 1:**
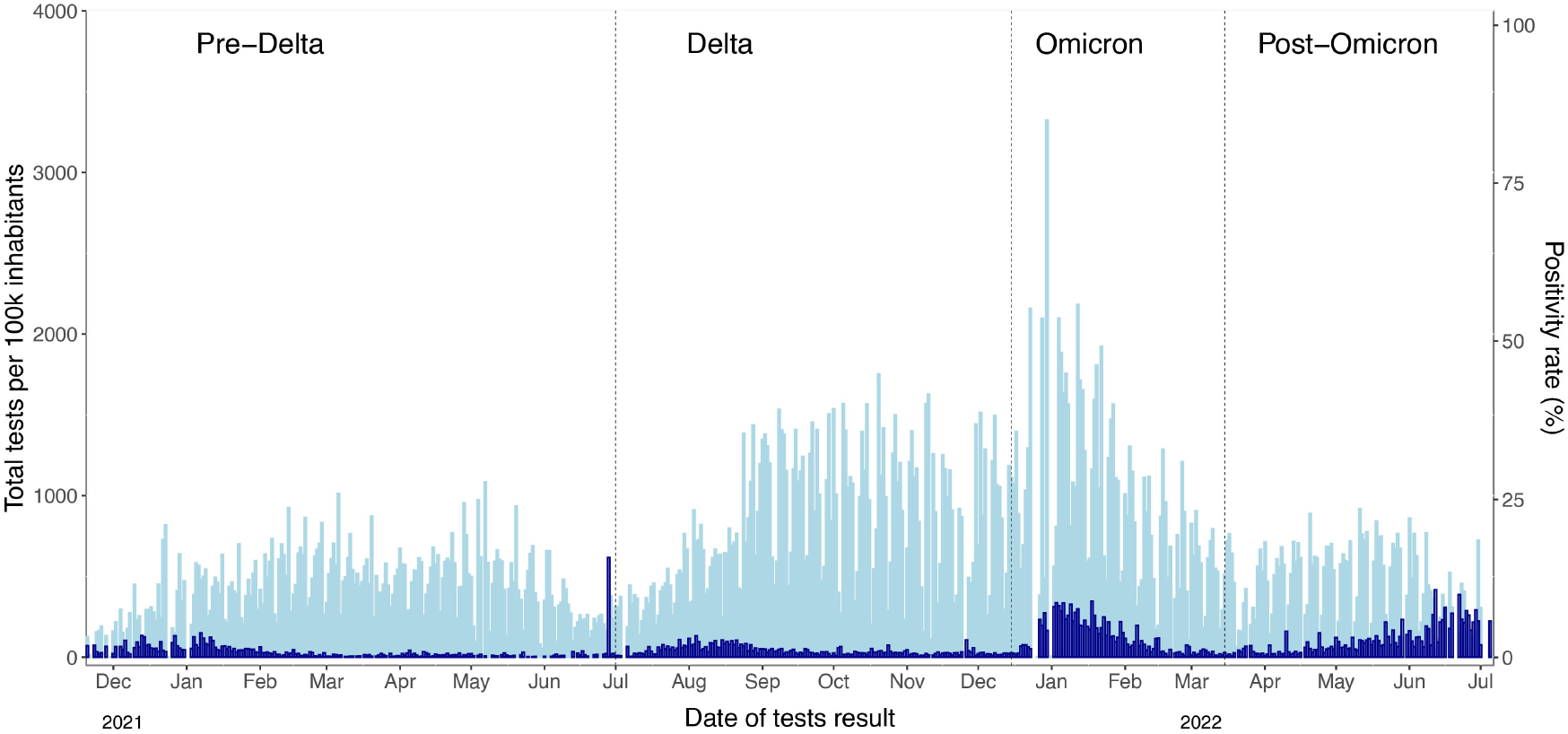
Daily tests performed by 100,000 population (light-blue) and daily positive rate (dark-blue) from November 18, 2020, to June 30, 2022 in Yolo County divided (dash-lines) into four variant periods.

This study was determined to be exempt from institutional review board review by the UC Davis Office of Research.

During the *HDT* program test participants were assigned a unique ID as one person might be tested several times throughout. Henceforth, we refer to *unique testers* as individuals that underwent a single test within each study period (e.g., *Omicron, Post-Omicron*). A participant may be a *unique tester* for a specific period (e.g., *Delta*), even if they underwent testing during a separate period (e.g., *Omicron, Post-Omicron*). Total tests, new tests, new testers and positive cases are described within each study period. Finally, we define a *frequent tester* as an individual that tested multiple times within a period of 15 days or less.

### 2.2 Tests Performed

Most tests conducted were PCR saliva tests (98.4%), with a small fraction of rapid antigen tests (1.6%). Saliva tests were based on reverse transcription – quantitative PCR test for asymptomatic individuals. BinaxNOW point-of-care tests were administered for symptomatic individuals. Saliva tests are non invasive and are more feasible for wide community implementation than antigen tests [29].

## 3 Results

HDT/HYT conducted 770,165 tests among 89,924 residents of Yolo County, of which 53,869 individuals reported a zip-code in the city of Davis. That is, the program reached approximately 41.1% of the population in Yolo County and 78.4% of Davis residents. Testing coverage was most significant in the City of Davis, where the program originated, and efforts were focused (See Figure A1). Figure 1 illustrates the total daily tests per 100,000 population and the positive rate observed in Yolo County throughout the study. The daily number of tests conducted per 100K during the *Pre-Delta* period was 422, *Delta* 746, *Omicron* 895, and *Post-Omicron* 435. Overall, the average number of total daily tests conducted was highest during *Omicron*, compared to other periods. A summary of tests and positive cases for each study period is reported in Table 1.

**Table 1:** Summary of testing by period. The numbers in parentheses reflect the rates in reference to the totals of the same category in the entire program.

| Description | Pre-Delta (208 days) |  |  | Delta (190 days) |  |  |
| --- | --- | --- | --- | --- | --- | --- |
|  | Total | Positive | Daily Av. | Total | Positive | Daily Av. |
| Total tests | 191904 (24.9%) | 1197 (9.5%) | 922.6 | 309928 (40.2%) | 2817 (22.3%) | 1631.2 |
| Unique testers | 39819 (44.3%) | 1138 (9.9%) | 191.4 | 58920 (65.5%) | 2577 (22.3%) | 310.1 |
| New testers | 39819 (44.3%) | 1138 (9.9%) | 191.4 | 33814 (37.6%) | 2570 (22.3%) | 178.0 |
|  | Omicron (85 days) |  |  | Post-Omicron (107 days) |  |  |
|  | Total | Positive | Daily Av. | Total | Positive | Daily Av. |
| Total tests | 166482 (21.6%) | 6351 (50.3%) | 1958.6 | 101851 (13.2%) | 2261 (17.9%) | 951.9 |
| Unique testers | 46545 (51.8%) | 5862 (50.8%) | 547.6 | 23566 (26.2%) | 2142 (18.6%) | 220.2 |
| New testers | 13503 (15.0%) | 5757 (49.9%) | 158.9 | 2788 (3.1%) | 2080 (18.0%) | 26.1 |

Total tests (N= 770,165); Unique testers (N=89,924) overall.
Total positive tests (12626); total individuals infected (N=11,545) overall.

Descriptive statistics of tests performed in Yolo showed that 48,652 (43.1%) participants were female and 39,519 (37.3%) were male. Test participation rates across all race/ethnic and age groups (*<*85 years) were at least 25%. The highest positivity rates were observed among American Indian/Alaska Native (7.6%), Native Hawaiian/Other Pacific Islander (5.6%), Latinos (4.5%), and people aged 20-44 years old (20-34: 6.5%; 35-44: 6.9%, Figure 2). The lowest participation rate in testing was observed in individual older than 85 years of age; this is not surprising as testing in the elderly population was mandatory at congregate living facilities for the elderly and supported by other state-level efforts to protect this vulnerable population.

**Figure 2:**
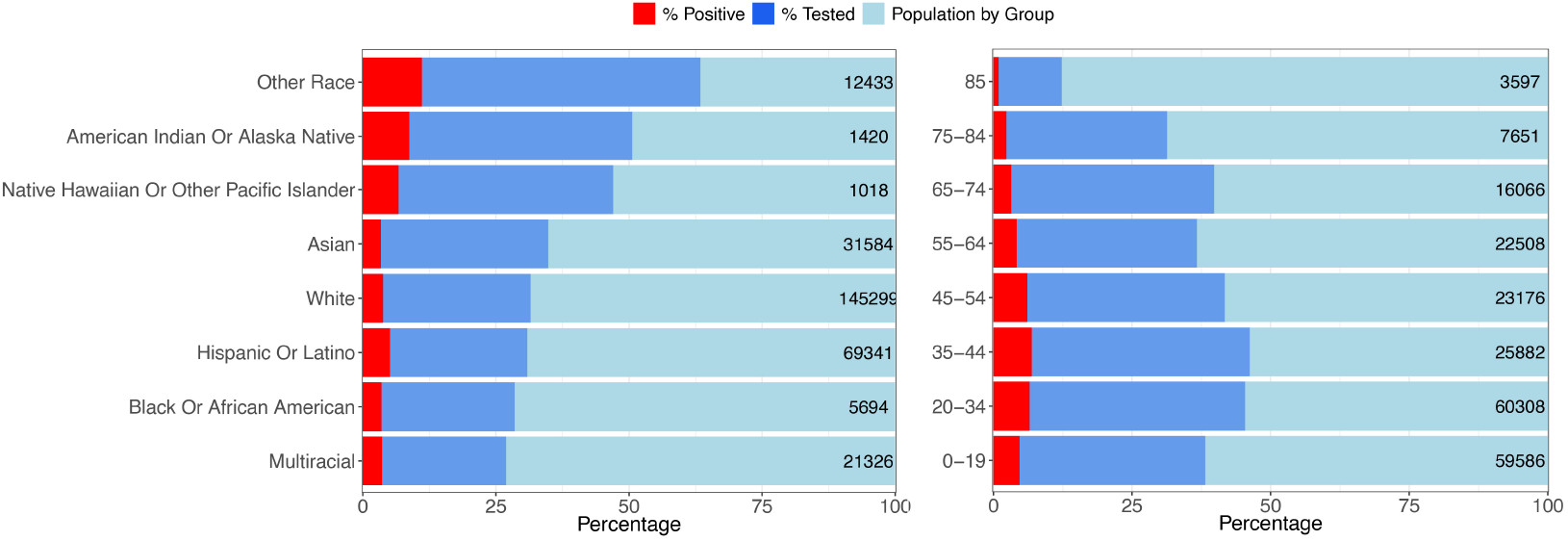
Tests and positive cases by race/ethnicity and age in years for Yolo County. Lightblue bars correspond to total population size for each group. Dark-blue bars correspond to the proportion of unique testers within each group; unique participants with a positive test results are depicted in red bars.

### 3.1 Testing participation over time

We describe the dynamics of test participation throughout the study in Figure 3 and Table 1. We testing of new and continuing participants varied widely over time. Overall, 12,626 tests were positive, among 11,545 individuals. Data presented in parenthesis of Table 1 correspond to estimates reflective of total population numbers. For instance, during *Pre-Delta* a total of 191,904 tests wee conducted, which corresponds to 24.9% of total tests (770,165) conducted in the entire program. During *Delta*, 2,570 positive cases were detected in new participants, corresponding to the 22.3% of the total of unique people resulted in positive (11,545).

**Figure 3:**
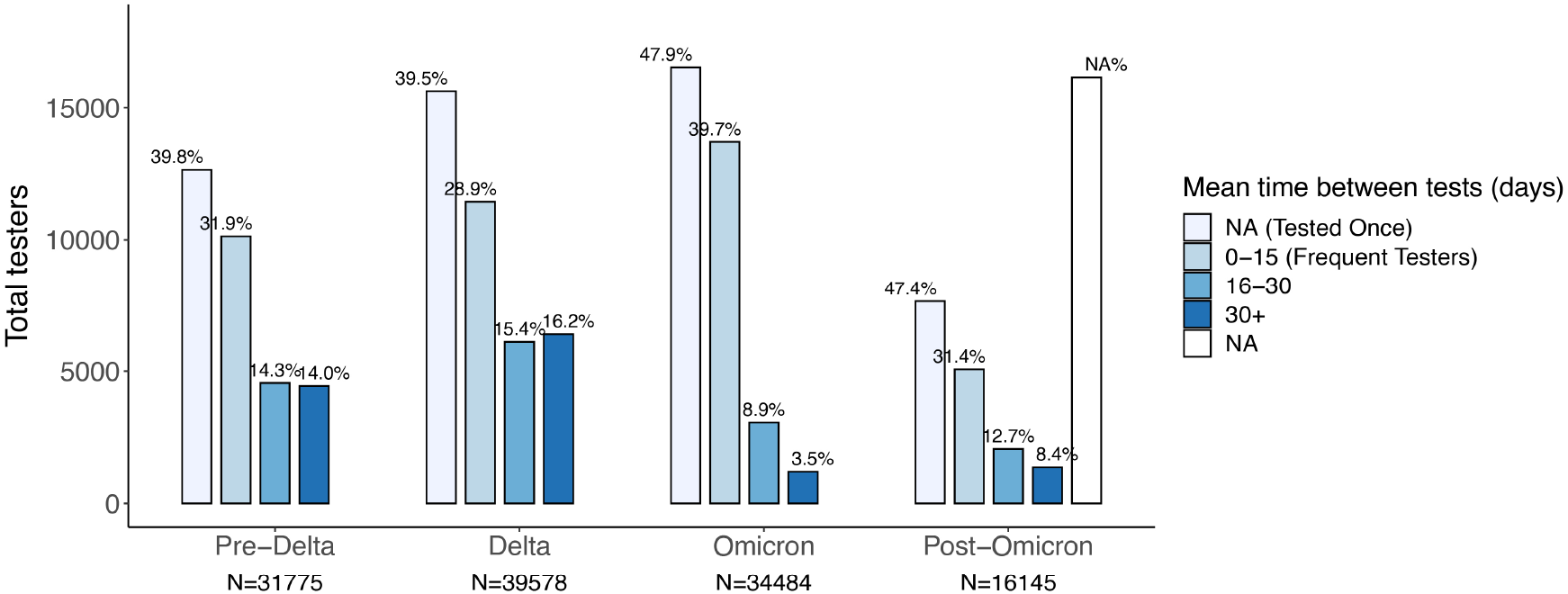
Changes in participation over time among non-student residents where *N* corresponds to unique testers. The bars represent the total number of individuals in four categories considering the time between tests (0-15 days, defined as frequent testers, 16-30 days, more than 30 days, and NA (not applicable) for people tested only once). Percentages represent the proportion of individuals tested among all tested within each per period.

The program was effective in engaging test participation throughout. In Yolo County, 39,819 (18.2%) individuals were tested in *Pre-Delta*, 58,920 (26.9%) during *Delta*, 46,545 (21.2%) in *Omicron*, and 23,566 (10.8%) in the *Post-Omicron* period. The *Omicron* period registered the highest daily rate of test participation 1959 daily average (∼ 895 per 100K), and highest number of positive cases (N=5,865, 50.8%) of all positive cases observed in the entire program. Participation from frequent testers was also most prominent during *Omicron* (Figure 3). Enrollment of new participants changed over time; 39,819 individuals were engaged in the first stage of the program, 33,814 new participants were tested during *Delta*, 13,503 during *Omicron*, and 2788 during the period of *Post-Omicron*.

Participation across different demographic groups remained steady (Table A1), with the exception of individuals under the age of 18 years (*Pre-Delta*: 20.2%, *Delta*: 32.8%, *Omicron*: 26.0%, *Post-Omicron*: 31.5%), with the lowest testing rate during *Pre-Delta*. Individuals self-identified as White (*Pre-Delta*: 58.9%, *Delta*: 52.9%, *Omicron*: 53.3%, *Post-Omicron*: 60.0%) experienced the highest rate of testing when compared to Latinos (*Pre-Delta*: 20.7%, *Delta*: 22.7%, *Omicron*: 21.9%, *Post-Omicron*: 16.4%).

### 3.2 Frequent testers

HDT/HYT collaborated with several school districts in Yolo County to provide weekly testing to students. Therefore, to identify the frequency of testing in the community, we excluded tests from school-age participants (*<*18 years, N=22,841). We classified the level of frequency in testing according to the average time between the tests performed by one person: 0-15 days (zero is assigned when an individual was tested multiple times on the same day), 16-30 days, more than 30 days, and NA (not applicable) indicates that people were tested only once.

Figure 3 illustrates the number of participants for each period according to the frequency of testing. The number and the proportion of frequent testers and people tested only once were higher during *Omicron* (Tables 2 and A2). The fraction of frequent testers increased significantly during *Omicron* (39.7%) when compared to *Pre-Delta* (31.9%), *Delta* (28.9%), and *Post-Omicron* (31.4%). The average time between tests in each period for frequent testers was between 6-7 days, and the maximum number of tests performed by one person was 116 in the *Delta* period (Figures A2-A3).

**Table 2:**
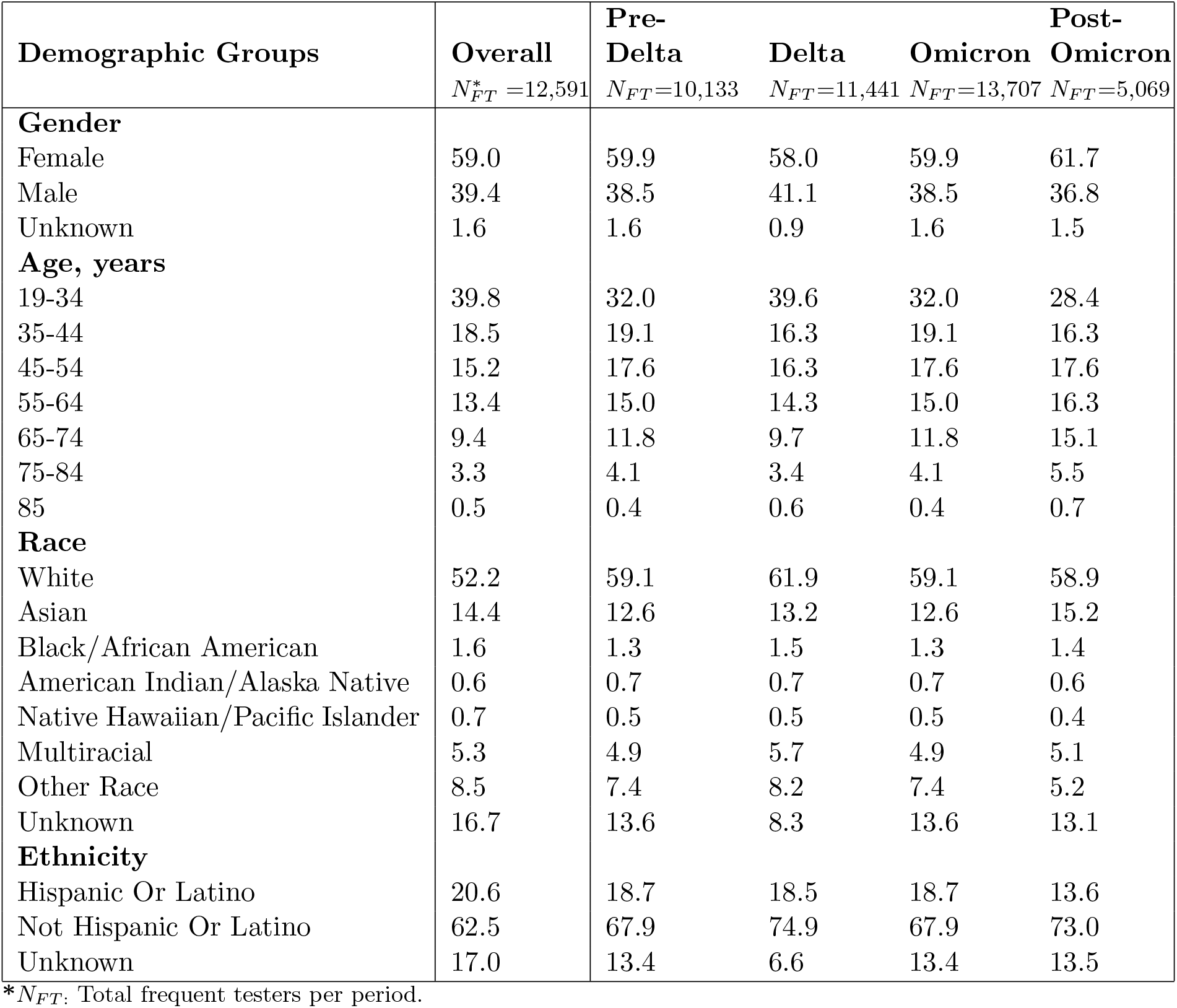
Demographic characteristics of frequent testers.

Table 2 summarizes the profile of frequent testers per period. Out of a total of 67,083 participants over the age of 18, approximately 18.8% (N=12,591) were tested regularly throughout the program. Among those frequent testers overall (N=12,591), the highest participation was registered by women (59%), people aged between 19-34 (39.8%), white people (52.2%), and Latino participation were around 20.6%. The highest number of frequent testers was recorded in the Omicron period, surpassing the number of frequent testers overall. The fact that a person was tested frequently during one period does not imply that they maintained that same practice throughout the program. The proportion of participation by demographic group remains similar over time. Women, individuals of age between 19-34, and White were consistently the most frequent testers in all study periods.

## 4 Discussion

This study describes the community asymptomatic testing experience in The Healthy Davis/Yolo Together (HDT/HYT) program, focusing on identifying participant demographics, changes in testing participation, and profiles of the most frequent testers. The HDT/HYT testing program had a high-throughput method for administering and processing large volumes of tests. It implemented testing strategies that allowed them to cover a significant percentage of the Davis and Yolo populations across all demographic groups who voluntarily underwent testing. In addition, testing campaigns and locations with free access to testing evolved throughout the program with the goal of reaching the most vulnerable or underserved.

Despite the empirical fact that men are more likely to experience adverse health consequences from COVID-19 [30, 31], a higher percentage of women than men participated in the program and were tested more frequently. These findings align with the existing literature documenting that women were more risk-averse than men during past infectious disease outbreaks [32, 33, 33] and that they are more favorable to adopting precautionary behaviors and following public health recommendations [34, 35, 36]. This finding is also consistent with other results that show women are more likely to report higher overall fear of coronavirus than males [37, 7]. Gender differences in risk perceptions may be due to deeply entrenched gender roles or differences in trusting authoritative figures and institutions [2]. Throughout the program, women, people ages 19-44, and whites were the most frequent raters.

User loyalty is a key factor in monitoring the spread and identifying the real burden of the disease. One of the objectives of epidemiological surveillance programs is to maintain user loyalty and enroll new users over time. Nevertheless, in a prolonged public health crisis such as COVID-19, pandemic fatigue is an expected and natural response [14]. Results show how effective the program was in retaining participants and engaging new people to participate in testing throughout the program. Omicron marks a turning point in the pandemic. It was the shortest period with the highest number of infections, the highest participation of individuals per day, and the one with the highest number of frequent testers. The increase in people participation and frequent testers may be related to the concern about the high risk of contagion given the growing number of cases or due to work, business, and school requirements. In return to normalcy, many places began to request COVID-19 negative tests or vaccination proofs to access services or keep work and school environments safe.

After Omicron period, people’s participation in testing dropped considerably. Motivation to engage in preventive behaviors against COVID-19 may decrease because people become tired and burn out due to excessive and repeated exposure to similar messages about COVID-19 over time [22, 15, 14]. Due to the impact of vaccines on risk perception, [22], the availability of other surveillance methods such as at-home COVID-19 tests [13], coupled with lack of trust in governments, misinformation, resistance, and conspiracy theories continue to pose a challenge for health authorities trying to maintain or revitalize public support to prevent COVID-19.

In long-term crises such as the COVID-19 pandemic, public health authorities may constantly face new challenges in maintaining preventive strategies due to changes in the virus and social behavior. The extensive data collected during the COVID-19 test campaigns provides an opportunity to learn about people’s adherence and practices towards public health measures, providing insights for the design of future strategies to educate communities about the benefits of engaging in preventive practices during new emerging infectious disease outbreaks. The HDT/HYT program offers a unique setting to analyze people’s behavior since non-compliance with prevention measures may be due to a reason other than limitations in access or availability of tests. But, the nature of our data do not allow us to conclude the reasons for the observed differences. However, patterns observed in this analysis comport with findings reported in previous studies with different with different data sources, such as survey information.

One of the great lessons the pandemic has taught us is the relevance of social dynamics in controlling the disease. Mathematical models were one of the most used tools to monitor the behavior of SARS-CoV-2. However, access to information that accounts for social behavior and people’s adherence to health policies was extremely limited as was access to granular population-based incidence data. Analysis of acceptance and response is a starting point not only to improve the design of future public health interventions but also to generate information that may be useful for designing of statistical and mathematical models to study the dynamics of infectious diseases.

This analysis looks at acceptance determinants using large-scale community PCR testing information. However, this kind of data allows only to identify socio-demographic information. To better understand what factors affected the uptake of free resources like testing or vaccine, it is necessary to collect data that captures individual behavioral characteristics for future studies to optimize the effectiveness of population-based interventions such as testing.

## Data Availability

All data produced in the present study are available upon reasonable request to the authors

