## Supplementary material for "Participant Demographics and Testing Trends: A Community Pandemic Response Program"

### 1 Participation over time by demographics

**Table A1.** Percentage of testers by demographic groups over time

| **Description** | **Overall** | **Pre-Delta** | **Delta** | **Omicron** | **Post-Omicron** |
|---|---|---|---|---|---|
| | $N^*$ =89,924 | $N$ =39,819 | $N$ =58,920 | $N$ =46,545 | $N$ =23,566 |
| **Gender** | | | | | |
| Female | 51.6 | 54.6 | 54.9 | 54.9 | 55.6 |
| Male | 40.3 | 43.9 | 42.9 | 43.1 | 42.0 |
| Unknown | 8.1 | 1.6 | 2.2 | 2.1 | 2.3 |
| **Age Group (years)** | | | | | |
| 0-18 | 31.6 | 20.2 | 32.8 | 26.0 | 31.5 |
| 19-34 | 19.2 | 32.8 | 23.6 | 27.3 | 20.6 |
| 35-44 | 10.7 | 12.8 | 13.5 | 13.7 | 12.6 |
| 45-54 | 10.6 | 11.8 | 10.8 | 11.6 | 11.5 |
| 55-64 | 9.9 | 10.4 | 8.8 | 9.9 | 10.9 |
| 65-74 | 8.5 | 8.2 | 7.2 | 8.1 | 9.3 |
| 75-84 | 3.1 | 3.1 | 2.8 | 3.0 | 3.2 |
| 85 | 0.4 | 0.6 | 0.4 | 0.4 | 0.4 |
| Unknown | 5.6 | - | - | - | - |
| **Race** | | | | | |
| White | 58.2 | 58.9 | 52.9 | 53.3 | 60.0 |
| Black Or African American | 1.3 | 1.7 | 1.6 | 1.6 | 1.2 |
| Asian | 12.1 | 12.0 | 10.8 | 12.2 | 11.2 |
| American Indian Or Alaska Native | 0.4 | 0.6 | 0.8 | 0.8 | 0.6 |
| Native Hawaiian Or Other Pacific Islander | 0.3 | 0.4 | 0.5 | 0.5 | 0.3 |
| Multiracial | 7.2 | 6.8 | 6.7 | 6.6 | 7.1 |
| Other Race | 4.3 | 8.5 | 7.6 | 8.0 | 5.5 |
| Unknown | 16.3 | 11.1 | 19.1 | 17.1 | 14.1 |
| **Ethnicity** | | | | | |
| Hispanic Or Latino | 11.9 | 20.7 | 22.7 | 21.9 | 16.4 |
| Not Hispanic Or Latino | 71.8 | 70.6 | 58.6 | 61.4 | 69.8 |
| Unknown | 16.3 | 8.7 | 18.7 | 16.7 | 13.8 |

$N$**:** Total unique testers per period.

### 2 Demographic coverage in the city of Davis

The Census Bureau 2020 may be biased respect to the demographic population in 2022. Thus, in some cases, comparing the number of people reported in the zip codes belonging to Davis exceeds the number projected by the data from the Census Bureau. In those cases, we assume a 100% coverage. Of 36,693 women in Davis, 29,002 (79.0%) were tested. Of 31,947 men, 23,815 (74.5%) were tested. As expected, the coverage in Davis was greater, more than 50% in all race/ethnicity and group ages (< 85).

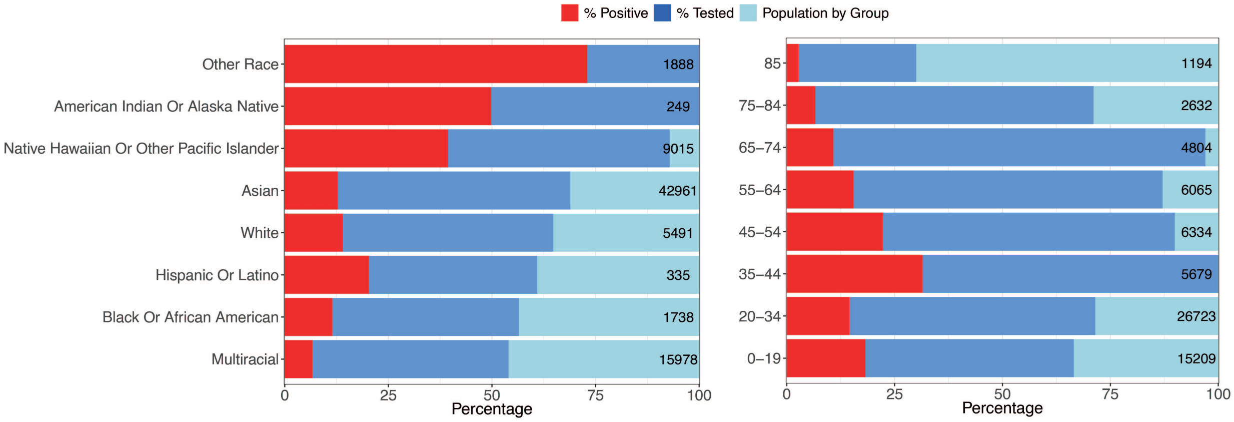


**Figure A1. Testing coverage and positive cases by demographics in the city of Davis** Light blue bars correspond to the total population by race/ethnicity. Dark blue bars correspond to the proportion of people the group covers, and Red bars correspond to unique positive cases.

### 3 Testing participation over time

#### Table A2. Testers different of students by frequency breakdown

| Mean time between tests (days) | Overall (N=67083 ) | | Pre-Delta (N=31775) | | Delta (N=39578) | | Omicron (N=34484) | | Post Omicron (N=16145) | |
|---|---|---|---|---|---|---|---|---|---|---|
| | Testers | Mean tests | Testers | Mean tests | Testers | Mean tests | Testers | Mean tests | Testers | Mean tests |
| Tested Ones | 23060 (34.4%) | 1 | 12650 (39.8%) | 1 | 15623(39.5%) | 1 | 16523 (47.9%) | 1 | 7654 (47.4%) | 1 |
| 0-15 | 12591 (18.8%) | 17 | 10133 (31.9%) | 9 | 11441 (28.9%) | 9 | 13707 (39.7%) | 5 | 5069 (31.4%) | 7 |
| 16-30 | 8403 (12.5%) | 13 | 4555 (14.3%) | 5 | 6111 (15.4%) | 5 | 3058 (8.9%) | 3 | 2058 (12.7%) | 4 |
| > 30 | 23029 (34.3%) | 5 | 4437 (13.9%) | 3 | 6403 (16.2%) | 3 | 1196 (3.5%) | 2 | 1364 (8.4%) | 2 |

[a] People tested only once.
**N:** Total participants per period.
(%) Proportion of tester in each category among total testers per period (N).
**Mean tests:** Average tests performed by person.

#### A4. Histograms of total tests and time between uptakes.

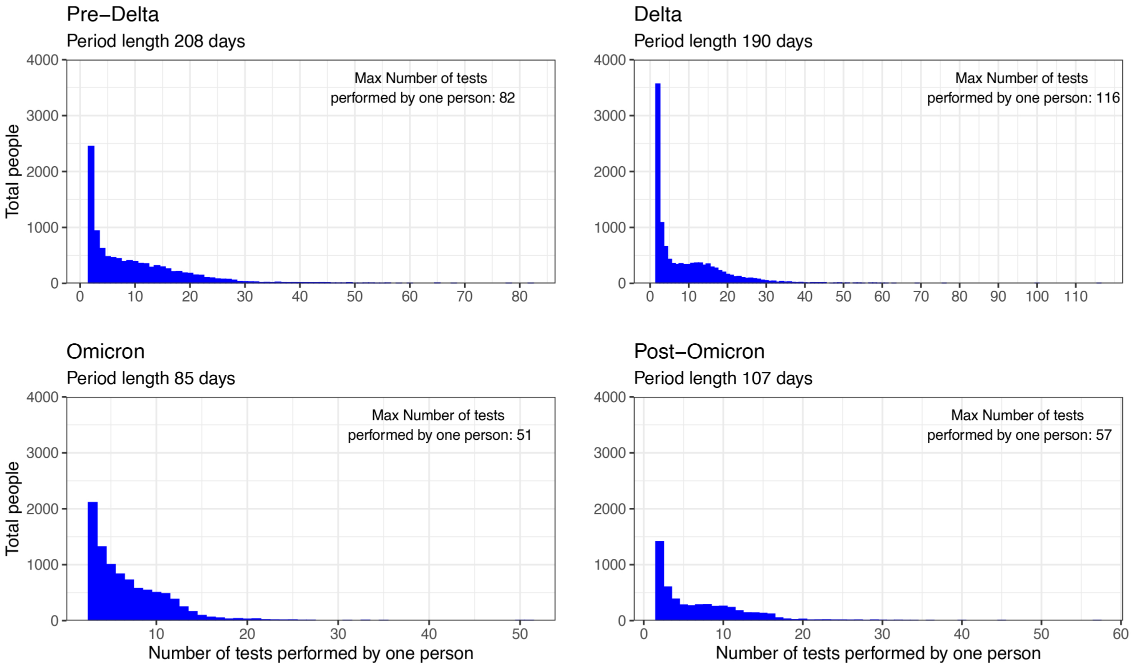


**Figure A2**. Distribution of the total number of tests carried out by participants.

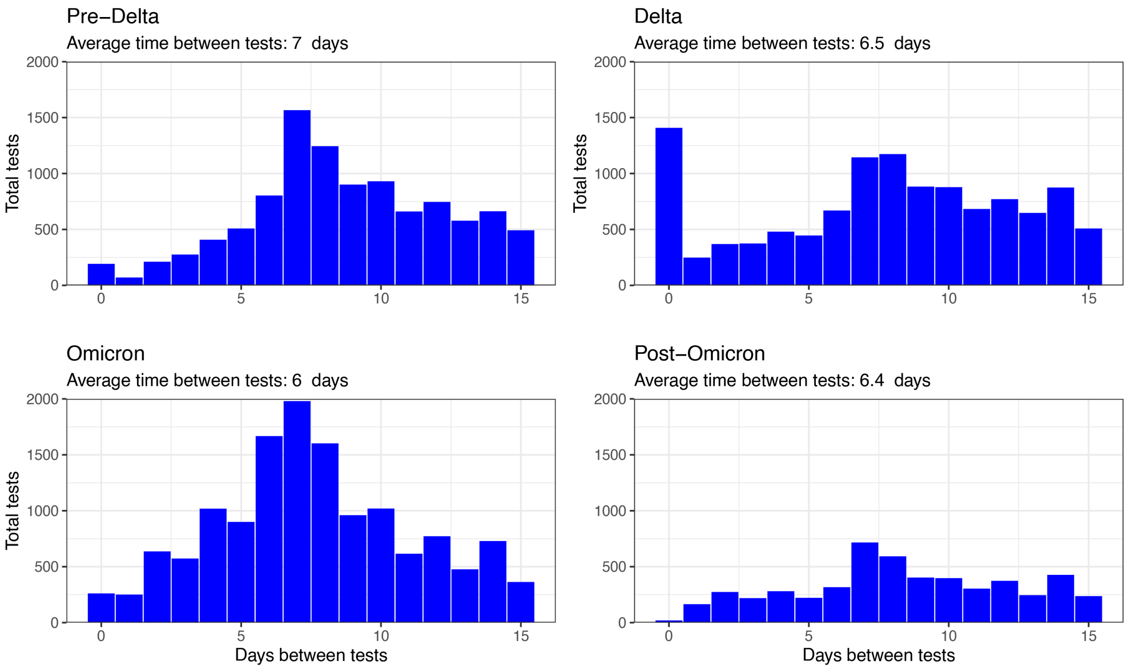


**Figure A3**. Distribution of days between tests carried aout by one participant.
